# Screening for rheumatoid arthritis-associated interstitial lung disease using lung ultrasound: a systematic review and meta-analysis of recent studies

**DOI:** 10.64898/2026.11.02.26365310

**Authors:** Armin Dan Copoeru, Mihaela Mocan, Anca Farcas, Sonia Vlaicu, Romeo Ioan Chira

## Abstract

Interstitial lung diseases (ILD) associated with systemic autoimmune disease can have a strong negative impact on patient outcomes. The diagnostic gold standard is high-resolution computer tomography (HRCT). Numerous studies have proven lung ultrasound's (LUS) utility in ILD, as well as other lung pathologies. Recently, there has been more focus on ILD in rheumatoid arthritis (RA), a previously overlooked complication, and screening methods. The purpose of this systematic review and meta-analysis was to summarize the recent knowledge regarding LUS compared to HRCT in screening ILD in RA patients. We reviewed relevant literature from the last 10 years and selected 12 articles. We included seven of those (totaling 756 patients) in a meta-analysis of diagnostic markers. The pooled sensitivity of LUS was 0.88 (95% CI 0.82-0.92) and pooled specificity was 0.86 (95% CI 0.71-0.94). The pooled LR+ and LR-were found to be 6.63 (95% CI 2.84-15.47) and 0.13 (95% CI 0.08-0.22), respectively. Diagnostic odds ratio was 48.54 (95% CI 13.78-170.98). LUS has high sensitivity and specificity for screening ILD in RA patients across different techniques and scoring systems.

## Introduction

Interstitial lung diseases (ILD) are a group of fibrotic diseases associated with diverse comorbidities, the most prevalent of which are systemic autoimmune diseases(1). The prototypical fibrosis-inducing autoimmune pathologies include systemic scleroderma, Sjogren’s syndrome and autoimmune myositis(2). Given that ILD are mostly clinically silent diseases with strong negative implications on quality of life and mortality, much research has gone into effective ways of diagnosis.(3–5) ILD are mainly classified based on morphological differences: usual interstitial pneumonia (UIP), which has the worst prognosis due to extensive tissue damage(6,7); non-specific interstitial pneumonia (NSIP), with a more favorable prognosis; other subtypes or unclassifiable ILD.(8)

The current gold standard is high-resolution computer tomography (HRCT), both for initial screening in patients with systemic autoimmune diseases, as well as later follow-up once diagnosis is established(9). It accurately detects, describes and quantifies fibrotic lung damage and has 30 years of research behind it. Nevertheless, it is a costly and irradiating procedure, thus limiting its utility in large-scale screening.

In light of this, research over the past decade has focused on alternate imagistic methods, of which the most promising is lung ultrasound (LUS)(10–18). While the lung was initially thought to be inaccessible to ultrasounds, it is now known that certain sonographic artefacts are closely associated with underlying fibrotic lung morphology and can accurately visualize ILD. B-lines, comet-tail artefacts that represent thickened interlobular septa, are the most important ultrasound abnormalities. They can be commonly seen alongside pleural line abnormalities (thickening and/or fragmentation)(19). The diagnostic opportunities afforded by LUS have been described in connective tissue diseases such as Sjogren’s syndrome (20), systemic sclerosis (21–25) and anti-synthetase syndrome(26,27). Furthermore, LUS is a much more accessible procedure insofar as screening is considered, and is non-irradiating.

Rheumatoid arthritis (RA) is a globally prevalent systemic autoimmune disease(28). It mainly affects small- and medium-sized joints, but is known to have a wide array of targets, including skin, eyes, muscle tissue, lungs and kidneys. Recent years have seen a shift of attention to lung fibrosis in RA (ILD-RA). Previously thought to be a rare complication, characteristic of other autoimmune diseases, it is now becoming increasingly evident that it has a higher prevalence and global burden.(29–32). It is also known that the most common morphological form of ILD-RA is UIP(33,34). Consequently, there is also increased interest in LUS’ diagnostic performance in ILD-RA(35,36).

The purpose of this systematic review is to summarize and evaluate the existing knowledge regarding sonographic screening accuracy in RA-ILD compared to the gold standard of HRCT.

## Methods

### Search strategy and article selection

We included in our search algorithm studies published in the last 10 years, up to 2016. The inclusion criteria were: diagnostic studies of LUS compared directly to HRCT in ILD in adult patients with confirmed RA. We excluded from this systematic review studies done on patients with other systemic autoimmune diseases or with mixed patient samples, other types of articles (case reports, study protocols etc.), those not available in English, articles with no available full text and animal studies.

This study was designed according to the 2015 PRISMA (Preferred Reporting Items for Systematic Review and Meta-Analysis) guidelines(37). A systematic search was performed in the electronic databases PubMed and Embase using the following combinations of MeSH keywords:

((“Ultrasonography”[Mesh]) AND “Arthritis, Rheumatoid”[Mesh]) AND “Pulmonary Fibrosis”[Mesh];

((“Ultrasonography”[Mesh]) AND “Lung Diseases, Interstitial”[Mesh]) AND “Arthritis, Rheumatoid”[Mesh];

((“Tomography, Spiral Computed”[Mesh]) AND “Arthritis, Rheumatoid”[Mesh]) AND “Pulmonary Fibrosis”[Mesh];

((“Tomography, Spiral Computed”[Mesh]) AND “Arthritis, Rheumatoid”[Mesh]) AND “Lung Diseases, Interstitial”[Mesh];

(“Ultrasonography”[Mesh]) AND “Pulmonary Fibrosis”[Mesh]; (“Lung Diseases, Interstitial”[Mesh]) AND “Ultrasonography”[Mesh]; (“Ultrasonography”[Mesh]) AND “Arthritis, Rheumatoid”[Mesh];

(“Arthritis, Rheumatoid”[Mesh]) AND “Lung Diseases, Interstitial”[Mesh].

After retrieving all studies, duplicates were removed and an initial analysis was done separately by two of the investigators by reviewing the studies’ titles and abstracts. Articles were either (a) excluded from the secondary analysis if they did not match the inclusion criteria or had at least one exclusion criterion or (b) included in the secondary analysis if they matched all of the inclusion criteria and had no exclusion criteria or did not have enough data in the title and abstract to definitely exclude. The investigators then confronted their findings; divergences were resolved by a third investigator. Secondary analysis was then performed by the two investigators by reviewing the studies fully, deciding whether to include them or not in this systematic review. Findings were again confronted, with divergences solved by a third investigator.

### Data extraction

After final article selection, data was organized based on the following topics: study design; patient inclusion & exclusion criteria; patient data (number, distribution based on age & sex, type of ILD); HRCT scoring method; LUS technique, scoring method, hardware used & settings recommendations; diagnostic results (sensitivity, specificity, positive & negative predictive value, positive & negative likelihood ratio, area under receiver operating characteristics curve) and other results. We also obtained true positive, false positive, true negative and false negative values, when available, from studies. Missing data points were noted as such.

### Quality assessment

Methodological quality assessment was done using the QUADAS-2 (Quality Assessment of Diagnostic Accuracy Studies) criterion to provide a standardized approach. QUADAS-2 classifies the risk of bias and study generalizability as high, low or unclear. Two independent investigators used the tool; in the case of a disagreement, a third investigator was consulted and discrepancies were resolved by consensus.

### Statistical analysis

First, we evaluated heterogeneity using the I^2^ statistic. When I^2^ was greater than 50, we interpreted it as significantly heterogeneous and used the random effects model; if it was under 50, we used the fixed effects model(38). Sensitivity, specificity, positive likelihood ratio (LR+), negative likelihood ratio (LR-), diagnostic odds ratio (DOR) and area under summary receiver-operating characteristics (SROC) curve were calculated from the pooled data. The 95% confidence interval (CI) was calculated for individual and pooled data. The software used was Meta-DiSc (version 2.0, Ramon y Cajal Hospital, Madrid, Spain)(39).

## Results

### Study selection

As shown in Fig. 1, a total of 2426 relevant articles were obtained through the search, of which 143 were duplicates. Initial analysis was done on 2283 articles and 2252 were excluded. The remaining 31 were assessed for eligibility. Finally, we identified 12 eligible studies for this systematic review, totaling 1141 patients(40–51). We excluded from the secondary analysis twenty studies: one animal study (52), one commentary (53), one article without available full text (54), one study with no HRCT comparison (55) and fifteen studies with mixed patient samples (56–70). Study design, patient number and details, inclusion & exclusion criteria and demographic data are shown in Table 1. HRCT scoring, LUS technique, scoring and hardware are presented in Table 2. The results of the studies are found in Table 3.

**Table 1.**
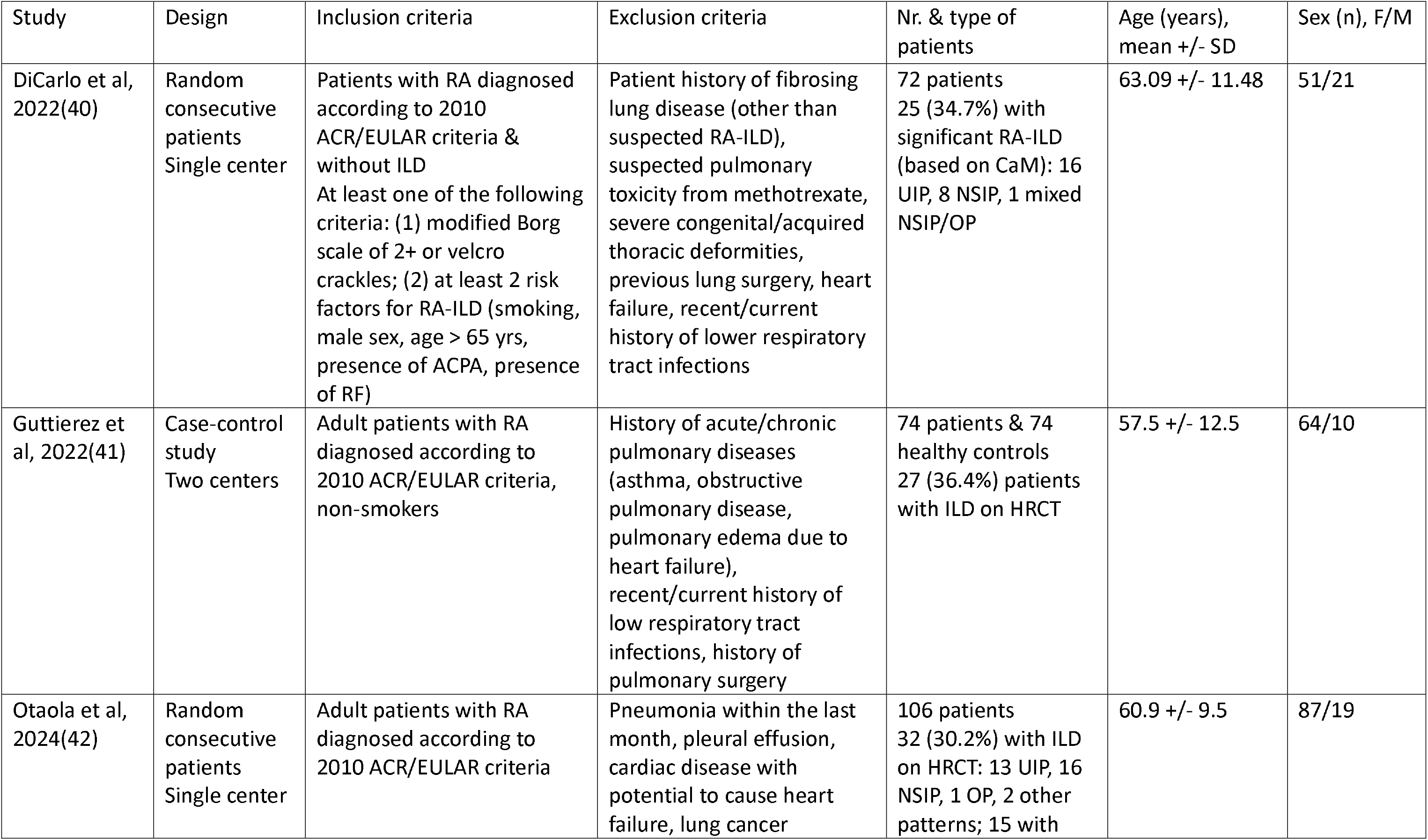
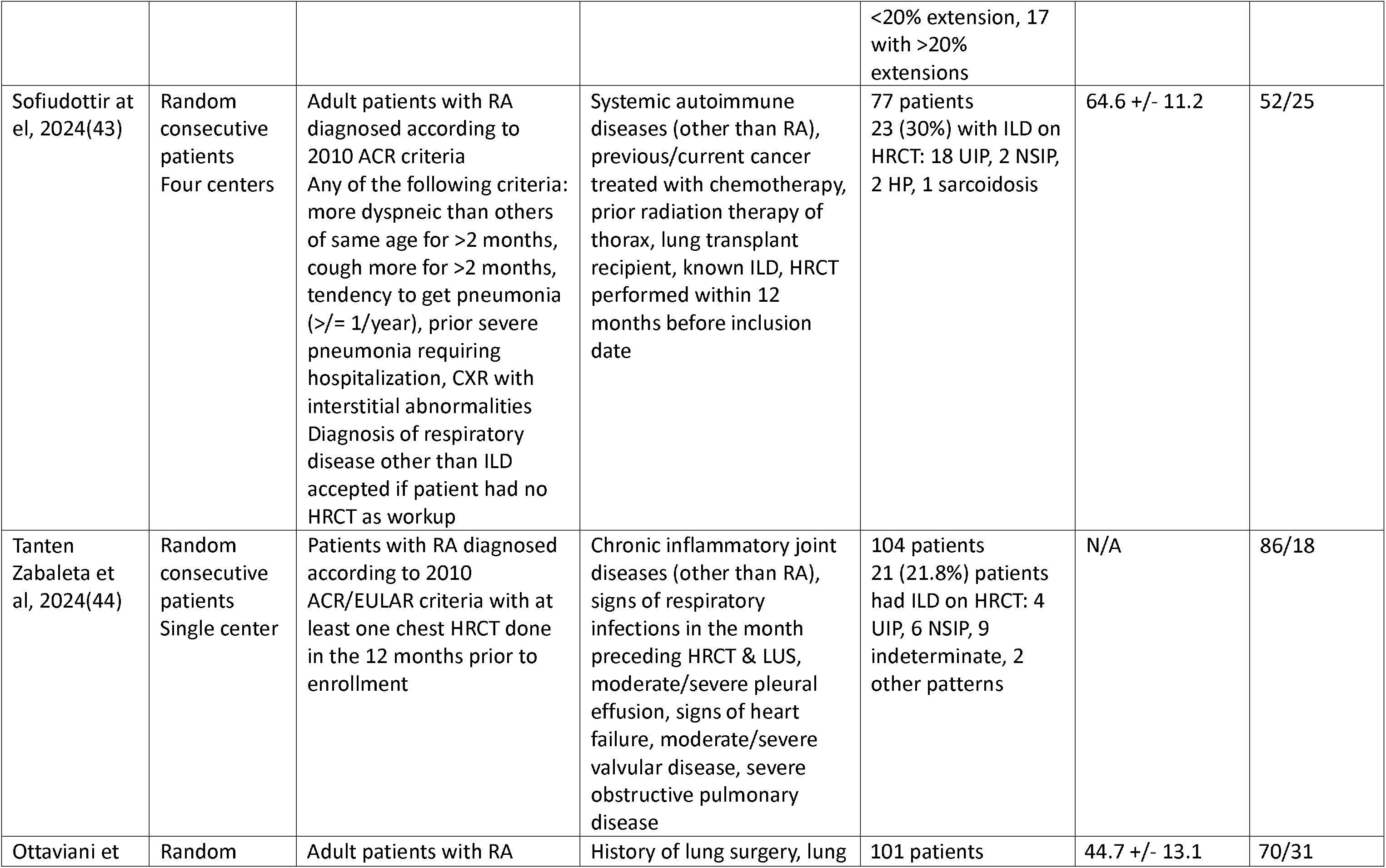
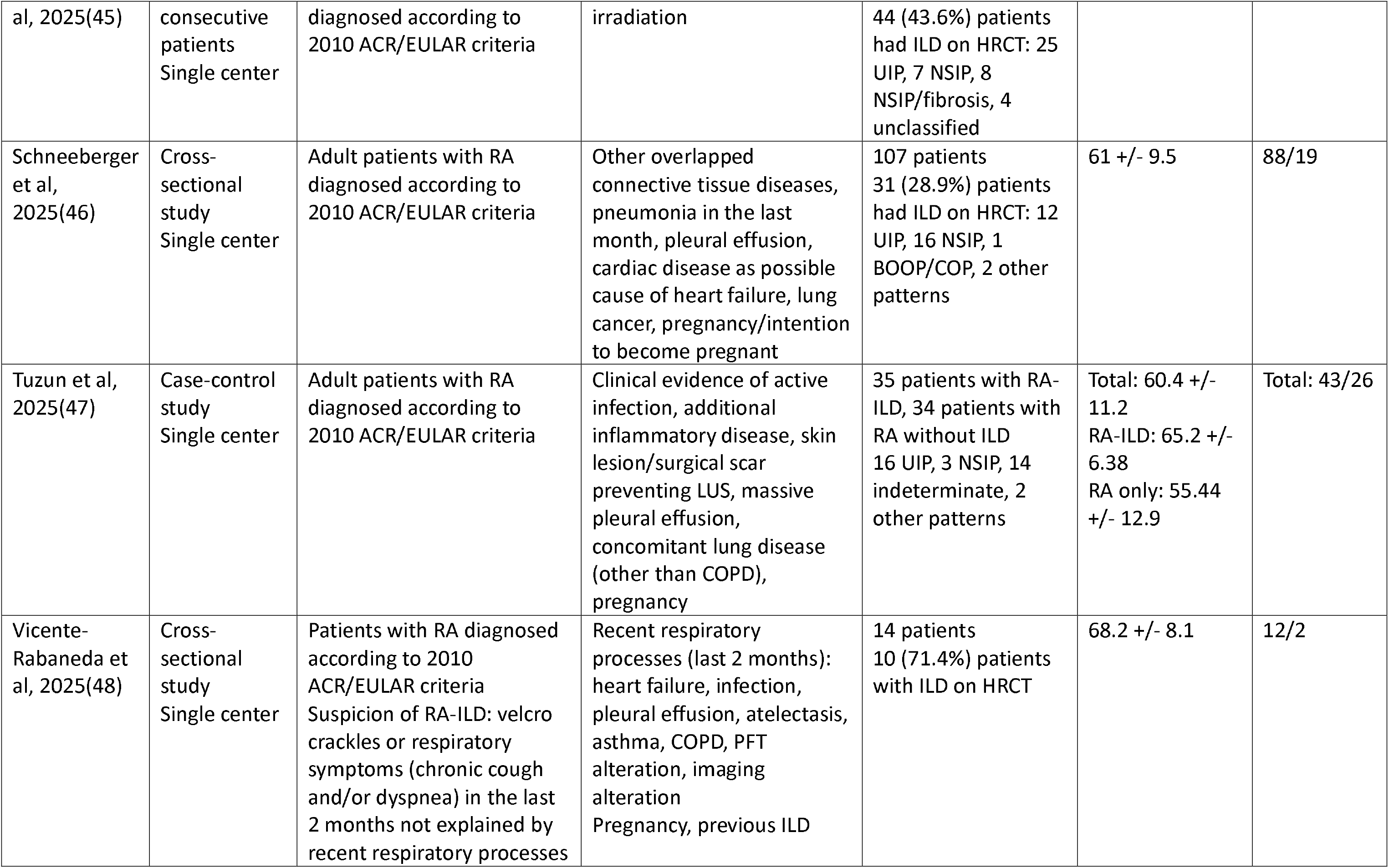
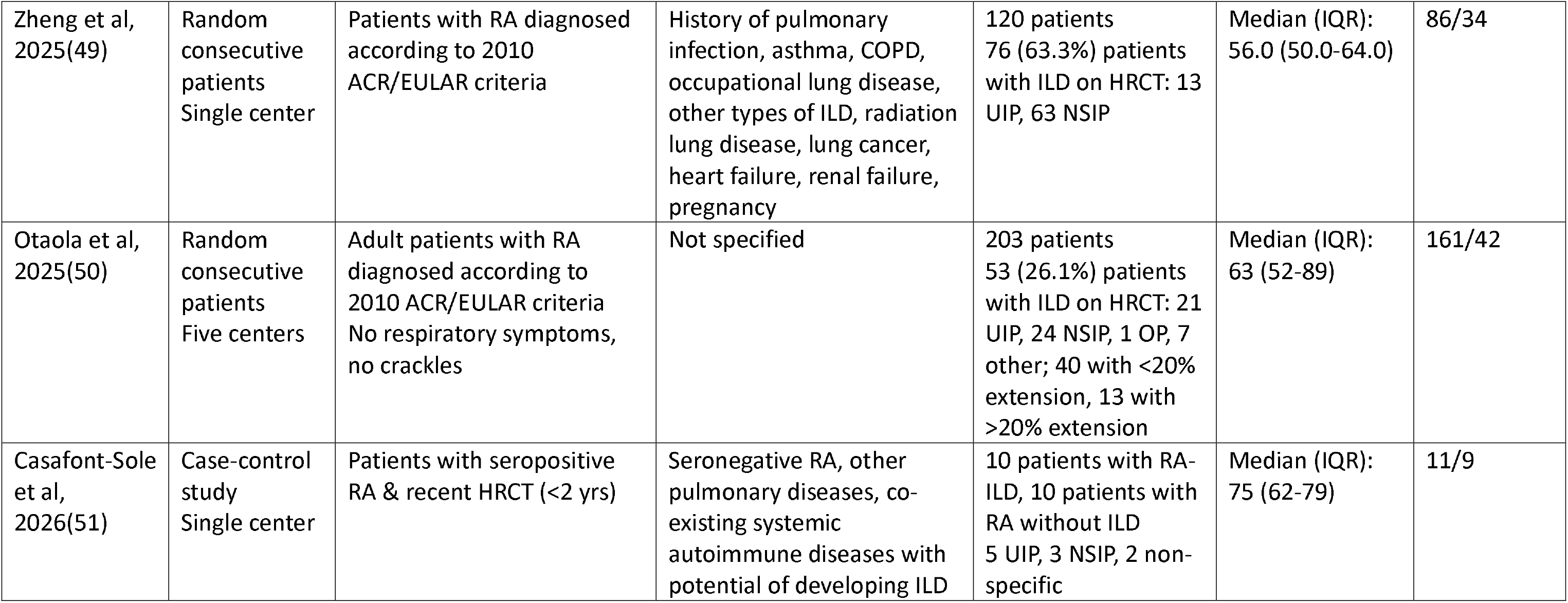
Study design, inclusion & exclusion criteria and patient data. ACPA=anti-citrullinated protein antibodies. ACR=American College of Rheumatology; BOOP=bronchiolitis obliterans organizing pneumonia; EULAR=European Alliance of Associations for Rheumatology; CaM=computer-aided method (see Table 2); COP=cryptogenic organizing pneumonia; COPD=chronic obstructive pulmonary disease; CXR=chest X-ray; HP=hypersensitivity pneumonitis; HRCT=high-resolution computer tomography; ILD=interstitial lung disease; LUS=lung ultrasound; NSIP=non-specific interstitial pneumonia; OP=organizing pneumonia; PFT=pulmonary function tests; RA=rheumatoid arthritis; RA-ILD=rheumatoid arthritis-associated interstitial lung disease; RF=rheumatoid factor; UIP=usual interstitial pneumonia.

| Study | Design | Inclusion criteria | Exclusion criteria | Nr. & type of patients | Age (years), mean +/- SD | Sex (n), F/M |
|---|---|---|---|---|---|---|
| DiCarlo et al, 2022(40) | Random consecutive patients<br>Single center | Patients with RA diagnosed according to 2010 ACR/EULAR criteria & without ILD<br>At least one of the following criteria: (1) modified Borg scale of 2+ or velcro crackles; (2) at least 2 risk factors for RA-ILD (smoking, male sex, age > 65 yrs, presence of ACPA, presence of RF) | Patient history of fibrosing lung disease (other than suspected RA-ILD), suspected pulmonary toxicity from methotrexate, severe congenital/acquired thoracic deformities, previous lung surgery, heart failure, recent/current history of lower respiratory tract infections | 72 patients<br>25 (34.7%) with significant RA-ILD (based on CaM): 16 UIP, 8 NSIP, 1 mixed NSIP/OP | 63.09 +/- 11.48 | 51/21 |
| Guttierez et al, 2022(41) | Case-control study<br>Two centers | Adult patients with RA diagnosed according to 2010 ACR/EULAR criteria, non-smokers | History of acute/chronic pulmonary diseases (asthma, obstructive pulmonary disease, pulmonary edema due to heart failure), recent/current history of low respiratory tract infections, history of pulmonary surgery | 74 patients & 74 healthy controls<br>27 (36.4%) patients with ILD on HRCT | 57.5 +/- 12.5 | 64/10 |
| Otaola et al, 2024(42) | Random consecutive patients<br>Single center | Adult patients with RA diagnosed according to 2010 ACR/EULAR criteria | Pneumonia within the last month, pleural effusion, cardiac disease with potential to cause heart failure, lung cancer | 106 patients<br>32 (30.2%) with ILD on HRCT: 13 UIP, 16 NSIP, 1 OP, 2 other patterns; 15 with <20% extension, 17 with >20% extensions | 60.9 +/- 9.5 | 87/19 |
| Sofiudottir at el, 2024(43) | Random consecutive patients<br>Four centers | Adult patients with RA diagnosed according to 2010 ACR criteria<br>Any of the following criteria: more dyspneic than others of same age for >2 months, cough more for >2 months, tendency to get pneumonia (>/= 1/year), prior severe pneumonia requiring hospitalization, CXR with interstitial abnormalities<br>Diagnosis of respiratory disease other than ILD accepted if patient had no HRCT as workup | Systemic autoimmune diseases (other than RA), previous/current cancer treated with chemotherapy, prior radiation therapy of thorax, lung transplant recipient, known ILD, HRCT performed within 12 months before inclusion date | 77 patients<br>23 (30%) with ILD on HRCT: 18 UIP, 2 NSIP, 2 HP, 1 sarcoidosis | 64.6 +/- 11.2 | 52/25 |
| Tanten Zabaleta et al, 2024(44) | Random consecutive patients<br>Single center | Patients with RA diagnosed according to 2010 ACR/EULAR criteria with at least one chest HRCT done in the 12 months prior to enrollment | Chronic inflammatory joint diseases (other than RA), signs of respiratory infections in the month preceding HRCT & LUS, moderate/severe pleural effusion, signs of heart failure, moderate/severe valvular disease, severe obstructive pulmonary disease | 104 patients<br>21 (21.8%) patients had ILD on HRCT: 4 UIP, 6 NSIP, 9 indeterminate, 2 other patterns | N/A | 86/18 |
| Ottaviani et al, 2025(45) | Random consecutive patients<br>Single center | Adult patients with RA diagnosed according to 2010 ACR/EULAR criteria | History of lung surgery, lung irradiation | 101 patients<br>44 (43.6%) patients had ILD on HRCT: 25 UIP, 7 NSIP, 8 NSIP/fibrosis, 4 unclassified | 44.7 +/- 13.1 | 70/31 |
| Schneeberger et al, 2025(46) | Cross-sectional study<br>Single center | Adult patients with RA diagnosed according to 2010 ACR/EULAR criteria | Other overlapped connective tissue diseases, pneumonia in the last month, pleural effusion, cardiac disease as possible cause of heart failure, lung cancer, pregnancy/intention to become pregnant | 107 patients<br>31 (28.9%) patients had ILD on HRCT: 12 UIP, 16 NSIP, 1 BOOP/COP, 2 other patterns | 61 +/- 9.5 | 88/19 |
| Tuzun et al, 2025(47) | Case-control study<br>Single center | Adult patients with RA diagnosed according to 2010 ACR/EULAR criteria | Clinical evidence of active infection, additional inflammatory disease, skin lesion/surgical scar preventing LUS, massive pleural effusion, concomitant lung disease (other than COPD), pregnancy | 35 patients with RA-ILD, 34 patients with RA without ILD<br>16 UIP, 3 NSIP, 14 indeterminate, 2 other patterns | Total: 60.4 +/- 11.2<br>RA-ILD: 65.2 +/- 6.38<br>RA only: 55.44 +/- 12.9 | Total: 43/26 |
| Vicente-Rabaneda et al, 2025(48) | Cross-sectional study<br>Single center | Patients with RA diagnosed according to 2010 ACR/EULAR criteria<br>Suspicion of RA-ILD: velcro crackles or respiratory symptoms (chronic cough and/or dyspnea) in the last 2 months not explained by recent respiratory processes | Recent respiratory processes (last 2 months): heart failure, infection, pleural effusion, atelectasis, asthma, COPD, PFT alteration, imaging alteration<br>Pregnancy, previous ILD | 14 patients<br>10 (71.4%) patients with ILD on HRCT | 68.2 +/- 8.1 | 12/2 |
| Zheng et al, 2025(49) | Random consecutive patients<br>Single center | Patients with RA diagnosed according to 2010 ACR/EULAR criteria | History of pulmonary infection, asthma, COPD, occupational lung disease, other types of ILD, radiation lung disease, lung cancer, heart failure, renal failure, pregnancy | 120 patients<br>76 (63.3%) patients with ILD on HRCT: 13 UIP, 63 NSIP | Median (IQR): 56.0 (50.0-64.0) | 86/34 |
| Otaola et al, 2025(50) | Random consecutive patients<br>Five centers | Adult patients with RA diagnosed according to 2010 ACR/EULAR criteria<br>No respiratory symptoms, no crackles | Not specified | 203 patients<br>53 (26.1%) patients with ILD on HRCT: 21 UIP, 24 NSIP, 1 OP, 7 other; 40 with <20% extension, 13 with >20% extension | Median (IQR): 63 (52-89) | 161/42 |
| Casafont-Sole et al, 2026(51) | Case-control study<br>Single center | Patients with seropositive RA & recent HRCT (<2 yrs) | Seronegative RA, other pulmonary diseases, co-existing systemic autoimmune diseases with potential of developing ILD | 10 patients with RA-ILD, 10 patients with RA without ILD<br>5 UIP, 3 NSIP, 2 non-specific | Median (IQR): 75 (62-79) | 11/9 |

**Table 2.** HRCT scoring and LUS scoring and technique. ALAT= Latin American Thoracic Society; ATS=American Thoracic Society; CaM=computer-aided method; ERS=European Respiratory Society; HRCT=High-resolution computer tomography; ILD=interstitial lung diseases; JRS= Japanese Respiratory Society.

| Study | HRCT scoring | LUS technique | LUS scoring | LUS device & probe | LUS settings |
|---|---|---|---|---|---|
| DiCarlo et al, 2022(40) | Warrick score<br>Computer-aided method (CaM): % of affected lung volume based on HU | 14 points technique (8th paravertebral, 8th subscapular, 8th posterior axillary line, 4th middle axillary line, 4th anterior axillary line, 4th midclavicular line, 2nd parasternal) | Total nr of B-lines (if confluent, counted as percentage of space occupied, divided by 10) | MyLab Class C, Easote<br>4-13 MHz linear probe | N/A |
| Guttierez et al, 2022(41) | Warrick score | 14 points technique (8th paravertebral, 8th subscapular, 8th posterior axillary line, 4th middle axillary line, 4th anterior axillary line, 4th midclavicular line, 2nd parasternal) | Semi-quantitative scoring: 0 pts=normal=0-5 B-lines; 1 pts=slight=6-15 B-lines; 2 pts=moderate=16-30 B-lines; 3 pts=severe=30+ B-lines | Esaote MyLab 70 XVG with 5-13 MHz broadband multifrequency linear probe<br>Marshall M15 with 4-12 MHz broadband multifrequency linear probe | N/A |
| Otaola et al, 2024(42) | Scans interpreted by experienced radiologist; in case of uncertainties, reviewed by another experienced radiologist<br>Dichotomous scoring: cut-off value of 20% diseased lung area | 14 points technique (8th paravertebral, 8th subscapular, 8th posterior axillary line, 4th middle axillary line, 4th anterior axillary line, 4th midclavicular line, 2nd parasternal) | Dichotomous scoring: cut-off value of 5 total B-lines | Mylab 70 XVG<br>1-8 MHz convex probe | N/A |
| Sofiudottir at el, 2024(43) | HRCTs with areas of ground-glass opacity, consolidation, reticulation, fibrosis with traction bronchiectasis, honeycombing, evident loss of volume were selected<br>Scans interpreted by ILD-specialized radiologist<br>"Yes" or "No" diagnosis | Straight-backed sitting positon<br>14 points technique based on Laursen et al (2 anterior points, 2 lateral points, 3 posterior points)(71,72) | ILD criteria: B-lines present in 2+ zones with 10+ total B-lines OR thickened and fragmented visceral pleura in 1+ zone bilaterally<br>"Yes" or "No" diagnosis | Hitachi Arietta V70<br>1-5 MHz convex probe (C251) | Lung preset, initial depth 10 cm, focus point 2 cm, initial gain 55 dB, compound imaging & tissue harmonics deactivated, 5 MHz frequency |
| Tanten Zabaleta et al, 2024(44) | Scans interpreted by ILD-expert pulmonologist<br>ILD defined as >10% lung involvement, regardless of pattern | 14 points technique (8th paravertebral, 8th subscapular, 8th posterior axillary line, 4th middle axillary line, 4th anterior axillary line, 4th midclavicular line, 2nd parasternal) | Total nr of B-lines<br>Total nr of spaces with pleural irregularities | Esaote Mylab class C<br>8-18 MHz linear probe & 4-8 MHz curved probe | N/A |
| Ottaviani et al, 2025(45) | Warrick score | Near-supine position for anterior & lateral chest, sitting position for posterior chest<br>14 points technique (8th paravertebral, 8th subscapular, 8th posterior axillary line, 4th middle axillary line, 4th anterior axillary line, 4th midclavicular line, 2nd parasternal) | Semi-quantitative scoring: 0 pts=normal=0-5 B-lines; 1 pts=slight=6-15 B-lines; 2 pts=moderate=16-30 B-lines; 3 pts=severe=30+ B-lines | Esaote MyLab70 or MyLabX8<br>5-13 MHz linear probe | N/A |
| Schneeberger et al, 2025(46) | Scans interpreted by experienced radiologist<br>ILD classification according to 2018 ATS/ERS/JRS/ALAT criteria | Supine position for anterior & lateral chest, sitting position for posterior chest<br>14 points technique (8th paravertebral, 8th subscapular, 8th posterior axillary line, 4th middle axillary line, 4th anterior axillary line, 4th midclavicular line, 2nd parasternal) | Total B-line score: for each area, pts=nr of B-lines if <5 B-lines; 5 pts if 5+ B-lines without white lung; 10 pts if white lung present<br>ILD criteria: score of 5+ total pts | Esaote Mylab 70 XVG<br>1-8 MHz convex probe | N/A |
| Tuzun et al, 2025(47) | Warrick score | Sitting or supine position<br>12 points technique (each hemithorax divided in anterior, lateral & posterior regions, then further divided into upper & lower zones) | cLUS score for each zone (0 pts=normal aeration=A-lines or 1-2 B-lines; 1 pts=moderate loss of aeration=3+ B-lines; 2 pts=severe loss of aeration=coalescent B-lines; 3 pts=complete loss of aeration=tissue-like pattern) | Esaote MyLab70 X-Vision<br>6-18 MHz linear probe | N/A |
| Vicente-Rabaneda et al, 2025(48) | N/A | Supine position for anterior & lateral chest, sitting position for posterior chest<br>14 points technique (8th paravertebral, 8th subscapular, 8th posterior axillary line, 4th middle axillary line, 4th anterior axillary line, 4th midclavicular line, 2nd parasternal) | Global B-line score: sum of all B-lines (if confluent, counted as percentage of space occupied, divided by 10); max. limit of 6 B-lines/space<br>Binary B-line score: for each space, 0 pts=0-2 B-lines, 1 pt=3+ B-lines<br>Semi-quantitative pleural line score: for each space, 0 pts=normal pleural line, 1 pt=mild alterations affecting <30% of pleural width, 2 pts=moderate alterations of 30-50% of pleural width, 3 pts=marked alterations and/or extension of >50% pleural width<br>Binary pleural line score: for each space, 0 pts=normal pleural line, 1 pt=any abnormality | Esaote MyLab Seven<br>3-13 MHz linear probe | Focus set at pleural line, depth of 6-8 cm for B-lines & 4-5 cm for pleural line, automatic image enhancements removed<br>Individual adjustments to improve image quality based on patient characteristics allowed |
| Zheng et al, 2025(49) | Scans interpreted by experienced radiologist<br>ILD classification according to 2013 ATS/ERS criteria | 50 points technique (after Gutierrez et al)(73) | Total nr of B-lines (if confluent, counted as percentage of space occupied, divided by 10) | Siemens Medical Solutions<br>2.5-3.5 MHz cardiac sector probe | Depth 18 cm, dynamic range 65 dB |
| Otaola et al, 2025(50) | Scans interpreted by experienced radiologist<br>ILD cases classified as <20% or >20% lung involvement | 14 points technique (8th paravertebral, 8th subscapular, 8th posterior axillary line, 4th middle axillary line, 4th anterior axillary line, 4th midclavicular line, 2nd parasternal) | Dichotomous scoring: cut-off value of 5 total B-lines | Esaote Mylab 70 XVG with convex (1-8 MHz) probe<br>Mindray M7 with convex probe<br>Esaote Mylab 70 or X8 with linear (4-12 MHz) probe<br>GE Versana Premier with linear (5-13 MHz) and convex (3-5 MHz) probes<br>Butterfly iQ with convex (1-10 MHz) probe | N/A |
| Casafont-Sole et al, 2026(51) | Scans interpreted by expert radiologist according<br>ILD classification according to 2023 ATS/ERS criteria | Seated & supine positions<br>14 points technique (8th paravertebral, 8th subscapular, 8th posterior axillary line, 4th middle axillary line, 4th anterior axillary line, 4th midclavicular line, 2nd parasternal) | B-line score (0; <3; 3-5; 5-10; >10 B-lines/field), pleural irregularities, pleural thickness | EUB-5500 Hitachi<br>Low & high frequency convex & linear probes | N/A |

**Table 3.**
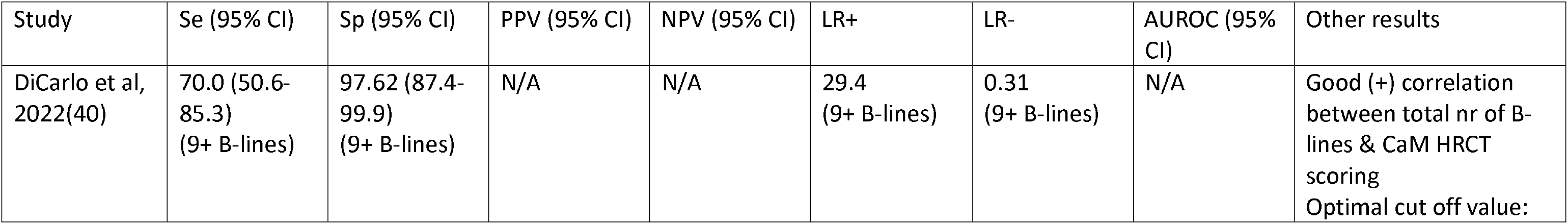
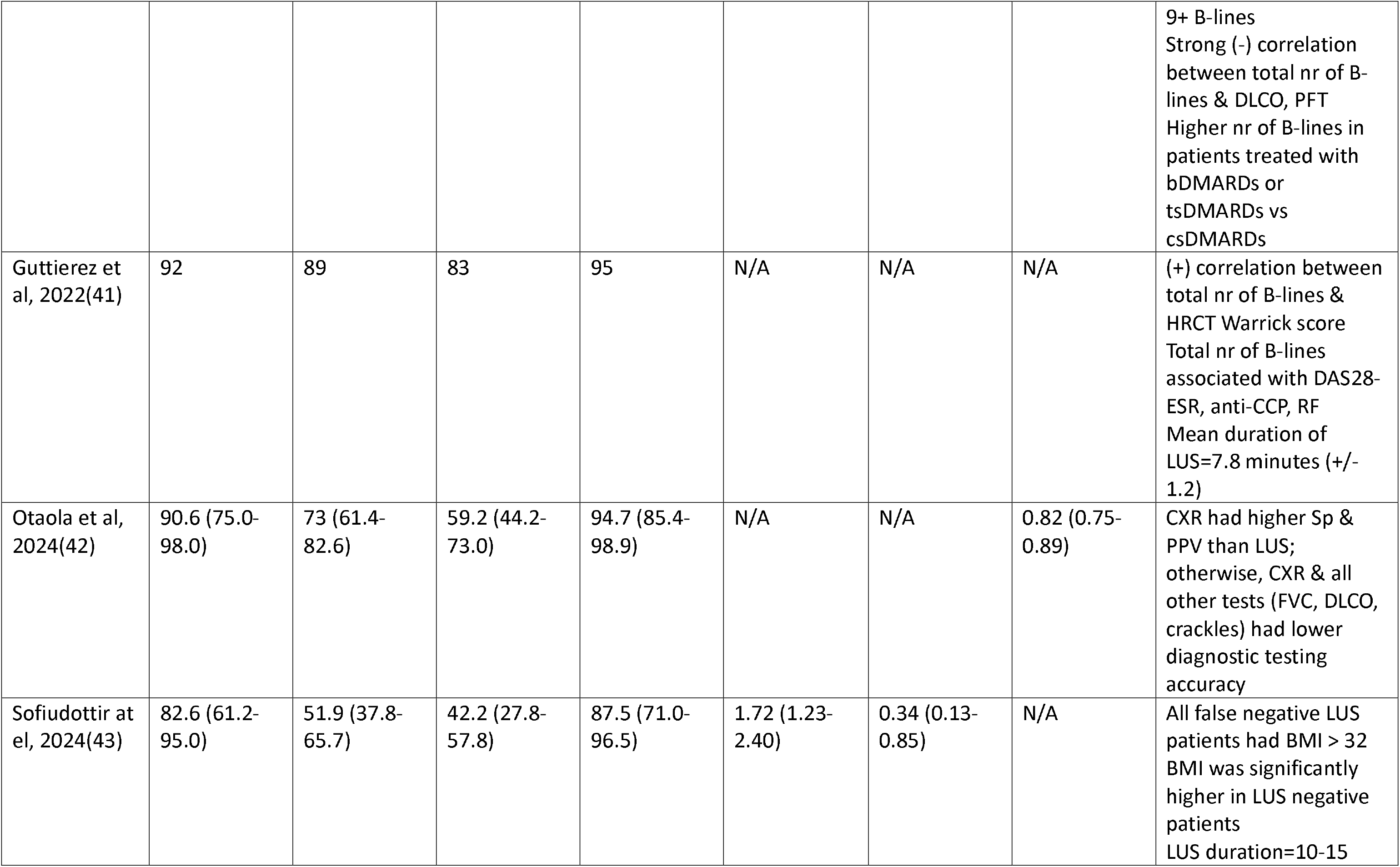
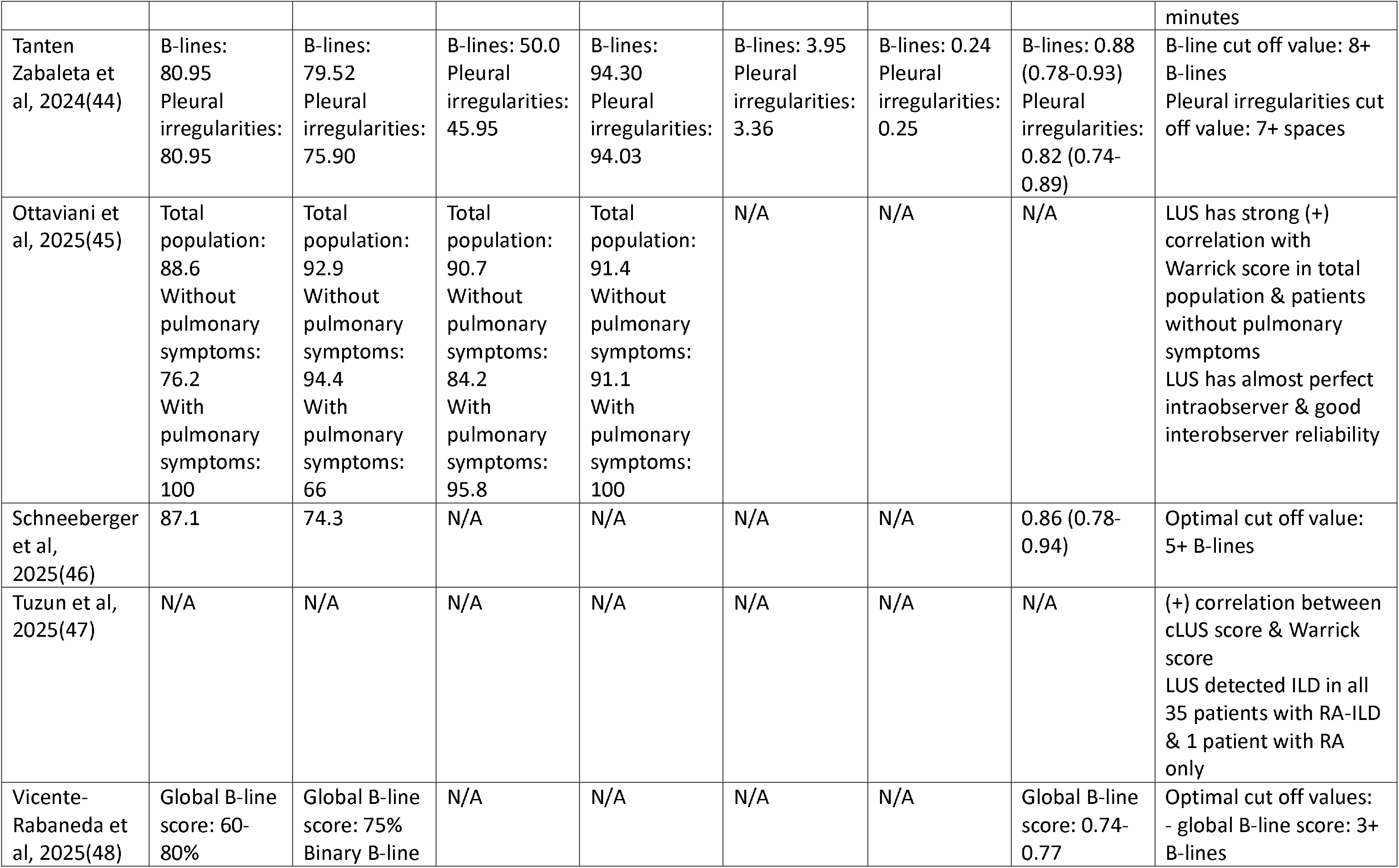
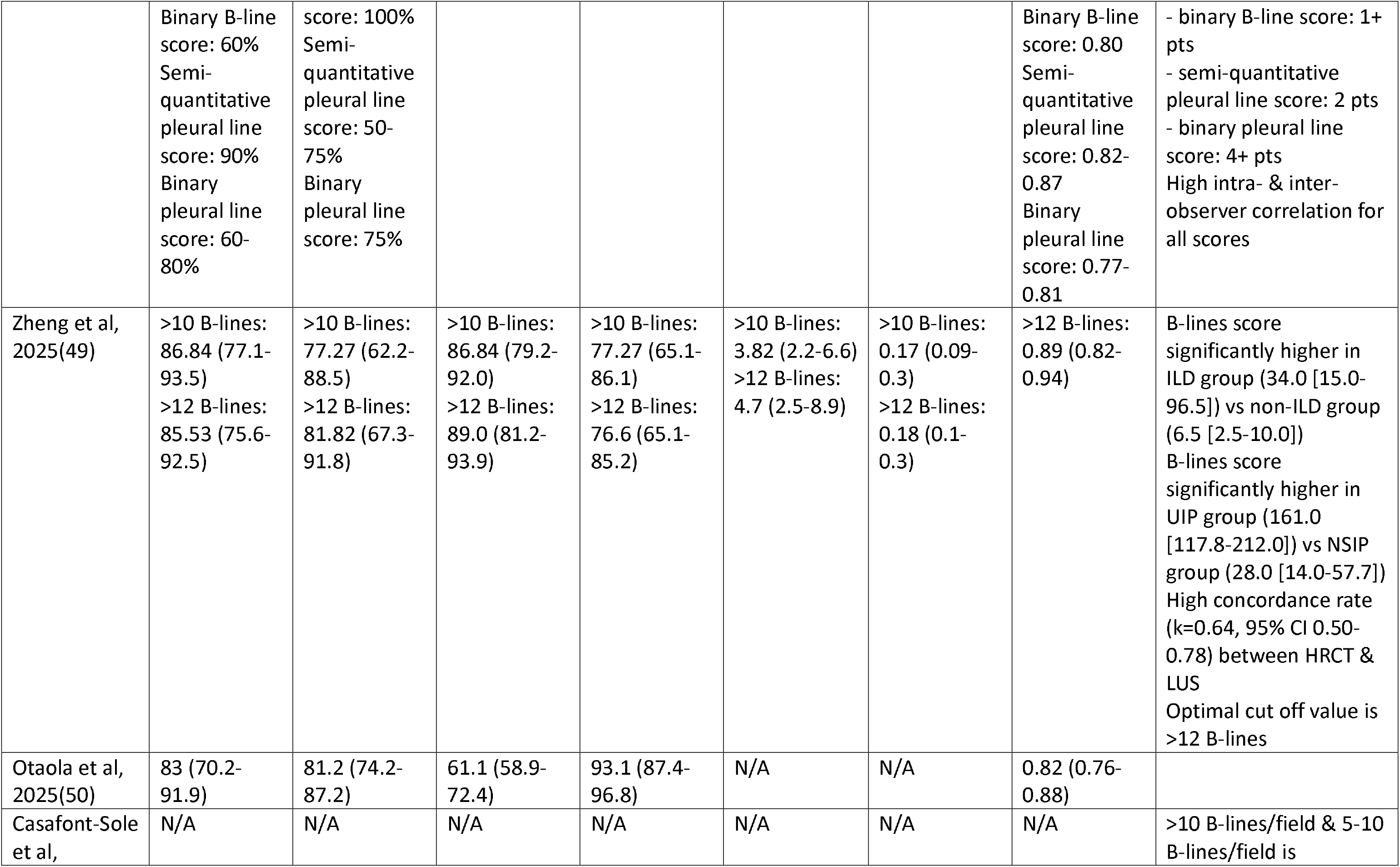
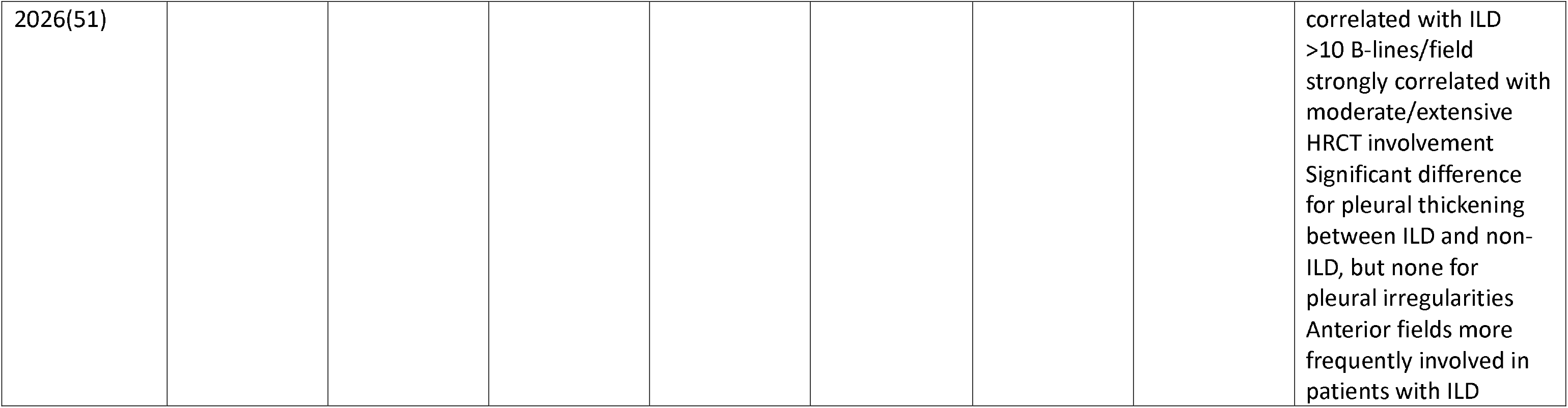
Diagnostic marker results. AUROC=area under receiver-operating characteristics curve; BMI=body-mass index; bDMARD=biological disease modifying anti-rheumatic drugs; CaM=computer-aided method; CCP=cyclic citrullinated peptide; csDMARD=conventional synthetic disease modifying anti-rheumatic drugs; CXR=chest X-ray; DAS28-ESR=disease activity score for rheumatoid arthritis with erythrocyte sedimentation rate; DLCO=diffusion capacity of carbon monoxide; FVC=forced vital capacity; HRCT=high-resolution computer tomography; ILD=interstitial lung disease; LR+=positive likelihood ratio; LR-=negative likelihood ratio; LUS=lung ultrasound; NPV=negative predictive value; NSIP=non-specific interstitial pneumonia; PPV=positive predictive value; RF=rheumatoid factor; Se=sensitivity; Sp=specificity; tsDMARD=targeted synthetic disease modifying anti-rheumatic drugs; UIP=usual interstitial pneumonia.

### Study characteristics

Seven studies enrolled patients in a random consecutive manner, three were case-control studies with either healthy subjects or RA patients without ILD and two studies were cross-sectional.

The lowest number of patients enrolled was 20 and the highest was 203; six studies enrolled more than 100 patients. Eleven studies sought patients with RA based on the 2010 ACR/EULAR criteria (four of which also had additional clinical & biological inclusion criteria), whilst one assessed patients with seropositive RA, with no mention of other diagnostic criteria. Most studies had effective and reasonable exclusion criteria, with one having limited exclusion criteria (history of lung surgery, lung irradiation) and another having no clearly specified exclusion criteria.

Regarding HRCT scoring, four studies used the Warrick score, seven classified ILD based on expert opinions or other severity scores, one gave no HRCT scoring method. The prevalence of ILD in RA patients ranged from 21.8% to 71.4%, with most patient samples being between 30-40%.

Ten studies used a 14-spaces LUS technique, one used a 50-spaces one and another used a 12-spaces one. The most used transducer was the linear one (eight studies), followed by convex probes (six studies) and cardiac sectorial probes (one study). Only three articles offered details about ultrasound settings. All articles evaluated B-lines and three studied pleural line abnormalities as well. Six studies used a semi-quantitative LUS scoring system, four had a quantitative score and four focused on a dichotomous score. Six articles had a pre-set LUS diagnostic threshold, while six had no initial thresholds. Two studies did not assess diagnostic accuracy markers.

### Meta-analysis

A total of 7 studies were eligible for meta-analysis based on available patient data (41–43,46,47,49,50), with a total of 756 patients, 281 with RA-ILD and 475 without (Table 4). For (49), we used only the patient data for the >10 B-lines threshold, as that was the pre-specified threshold. Heterogeneity was found to be high (see Supplemental file 1).

**Table 4.** Diagnostic data for studies included in meta-analysis.

| Study | True positive rate | False positive rate | True negative rate | False negative rate |
|---|---|---|---|---|
| Guttierez, 2022(41) | 25 | 2 | 42 | 5 |
| Otaola, 2024(42) | 29 | 20 | 54 | 3 |
| Sofiudottir, 2024(43) | 19 | 26 | 28 | 4 |
| Schneeberger, 2025(46) | 28 | 3 | 73 | 3 |
| Tuzun, 2025(47) | 35 | 1 | 33 | 0 |
| Zheng, 2025(49) | 66 | 10 | 34 | 10 |
| Otaola, 2025(50) | 44 | 28 | 121 | 10 |

The pooled sensitivity and specificity of LUS were 0.88 (95% CI 0.82-0.92) and 0.86 (95% CI 0.71-0.94), respectively (Fig. 2 and 3). The pooled LR+ and LR-were found to be 6.63 (95% CI 2.84-15.47) and 0.13 (95% CI 0.08-0.22), respectively (Table 5). DOR was 48.54 (95% CI 13.78-170.98) (Table 5). SROC curve is shown in Fig. 4.

**Table 5.** Pooled LR+, LR- and DOR values.

|  | Estimate | Lower limit of 95% CI | Upper limit of 95% CI |
|---|---|---|---|
| Sensitivity | 0.882 | 0.824 | 0.922 |
| Specificity | 0.867 | 0.718 | 0.944 |
| DOR | 48.544 | 13.781 | 170.988 |
| LR+ | 6.632 | 2.843 | 15.474 |
| LR- | 0.137 | 0.084 | 0.223 |

### Bias risk

The QUADAS-2 assessment is detailed in Table 6. The included studies’ overall quality was acceptable. Most studies enrolled patients with RA based on current diagnostic guidelines and had valid exclusion criteria. Five studies did not use a random consecutive sampling method, three of which opted for case-control design. HRCT was generally performed in an acceptable time frame before or after LUS, only one study accepting up to 2 years old HRCTs. Several studies were unclear about the HRCT and/or LUS operators’ blinding.

**Table 6.** QUADAS-2 assessment of included studies.

| Study | Risk of bias |  |  |  | Applicability concerns |  |  |
|---|---|---|---|---|---|---|---|
|  | Patient selection | Index test | Reference standard | Flow & timing | Patient selection | Index test | Reference standard |
| DiCarlo et al, 2022(40) | Low | Low | Low | Low | Low | Low | Low |
| Gutierrez et al, 2022(41) | High | Low | Low | Low | High | Low | Low |
| Otaola et al, 2024(42) | Unclear | Low | Unclear | Low | Unclear | Low | Unclear |
| Sofiudottir et al, 2024(43) | Low | Low | Low | Low | Low | Low | Low |
| Tanten Zabaleta et al, 2024(44) | Low | Unclear | High | Low | Low | Unclear | High |
| Ottaviani et al, 2025(45) | Unclear | Low | Low | Low | Unclear | Low | Low |
| Schneeberger et al, 2025(46) | Unclear | Low | Unclear | Low | Unclear | Low | Unclear |
| Tuzun et al, 2025(47) | High | Low | Unclear | Unclear | High | Low | Unclear |
| Vicente-Rabaneda et al, 2025(48) | Unclear | Low | High | Unclear | Unclear | Low | High |
| Zheng et al, 2025(49) | Low | Low | Unclear | Low | Low | Low | Unclear |
| Otaola et al, 2025(50) | Unclear | Low | Low | Low | Low | Low | Low |
| Casafont-Sole et al, 2026(51) | High | Low | Unclear | Low | High | Low | Unclear |

## Discussion

We found that LUS had high diagnostic accuracy across all included studies for the diagnosis of ILD-RA in adults. Results were consistently good across various diagnostic and prognostic scores and compared to different HRCT scoring techniques. This highlights ultrasound’s flexibility and reliability in diagnosing interstitial lung diseases. The main diagnostic elements for ILD-RA were number of B-lines and pleural abnormalities (thickening and/or irregularity). Most studies had clear and reasonable exclusion criteria (other ILDs, other acute or chronic lung diseases, heart failure etc.), which allowed for a stricter assessment of ultrasound’s capabilities. Real-life clinical practice may see lower diagnostic accuracy due to overlapping cardio-pulmonary diseases, which warrants further study and personnel training to avoid diagnostic pitfalls.

Sofiudottir et al (43) showed lower specificity and PPV than other studies due to a higher number of sonographic false-negatives. The authors associated the false-negatives with a BMI over 32, given that increased thoracic adipose tissue can independently reduce the number of B-lines(74). Furthermore, the diagnostic discrepancy might be explained by the study’s stricter binary diagnostic criteria, which might have reduced the number of true-positives.

LUS compared to HRCT fro RA-ILD was accurate according to the meta-analysis. It consistently proved to be adequately sensitive and specific, with a tendency towards a higher number of false positive diagnoses compared to false negatives. This could be at least partly explained by similar ultrasonographic aspects of ILD with other lung diseases (such as interstitial pneumonia or cardiogenic interstitial edema) and ultrasound’s tendency to miss earlier stages of fibrosis. Nevertheless, this tendency towards more false positives does not impede ultrasound’s use as an accessible screening tool.

To the best of our knowledge, this meta-analysis was the first one performed on RA-ILD comparing LUS to HRCT.

The results were compatible with those of studies focused on other systemic autoimmune diseases or those with mixed patient samples (75–78). Thus, LUS is equally efficient for RA patients as for those with other connective tissue diseases.

Most included studies showed a high prevalence of ILD in RA patients (between 20-40%). This finding contrasts with recent populational studies on ILD-RA(79), which showed a much lower incidence. This discordance is partly explained by some studies’ more specific inclusion criteria (previous suspicion of ILD based on clinical and/or radiological characteristics). On the other hand, high incidence was present in some studies with no other inclusion criteria besides confirmed RA diagnosis. Otaola et al(50) showed a prevalence of 26.1% in a group of asymptomatic RA patients. It is becoming increasingly clear that ILD-RA is an underestimated and underdiagnosed complication. Physicians should have raised awareness of this disease.

This systematic review and meta-analysis had some shortcomings of its own. First, the number of studies and patients included was small, as we focused on fibrotic complications from a single connective tissue disease; this can affect results and prevalence rates. Second, as mentioned above, almost all studies had extensive exclusion criteria; real-world use of LUS might thus show lower diagnostic rates when factoring in confounding factors such as other concomitant pulmonary diseases. Third, there was a great amount of methodological heterogeneity among the studies. Few used the same HRCT and LUS scoring system and sonographic hardware varied between all three types of probes.

## Conclusion

Lung ultrasound proves itself to be an accurate and flexible screening procedure for ILD-RA. Diagnostic accuracy markers are high across a wide spectrum of protocols, scores and patient populations. Future efforts should focus on systematic evaluation and long-term follow-up. Medical practitioners should have increased awareness of ILD-RA and its prognostic impact.

## Data Availability

All data produced in the present work are contained in the manuscript

---

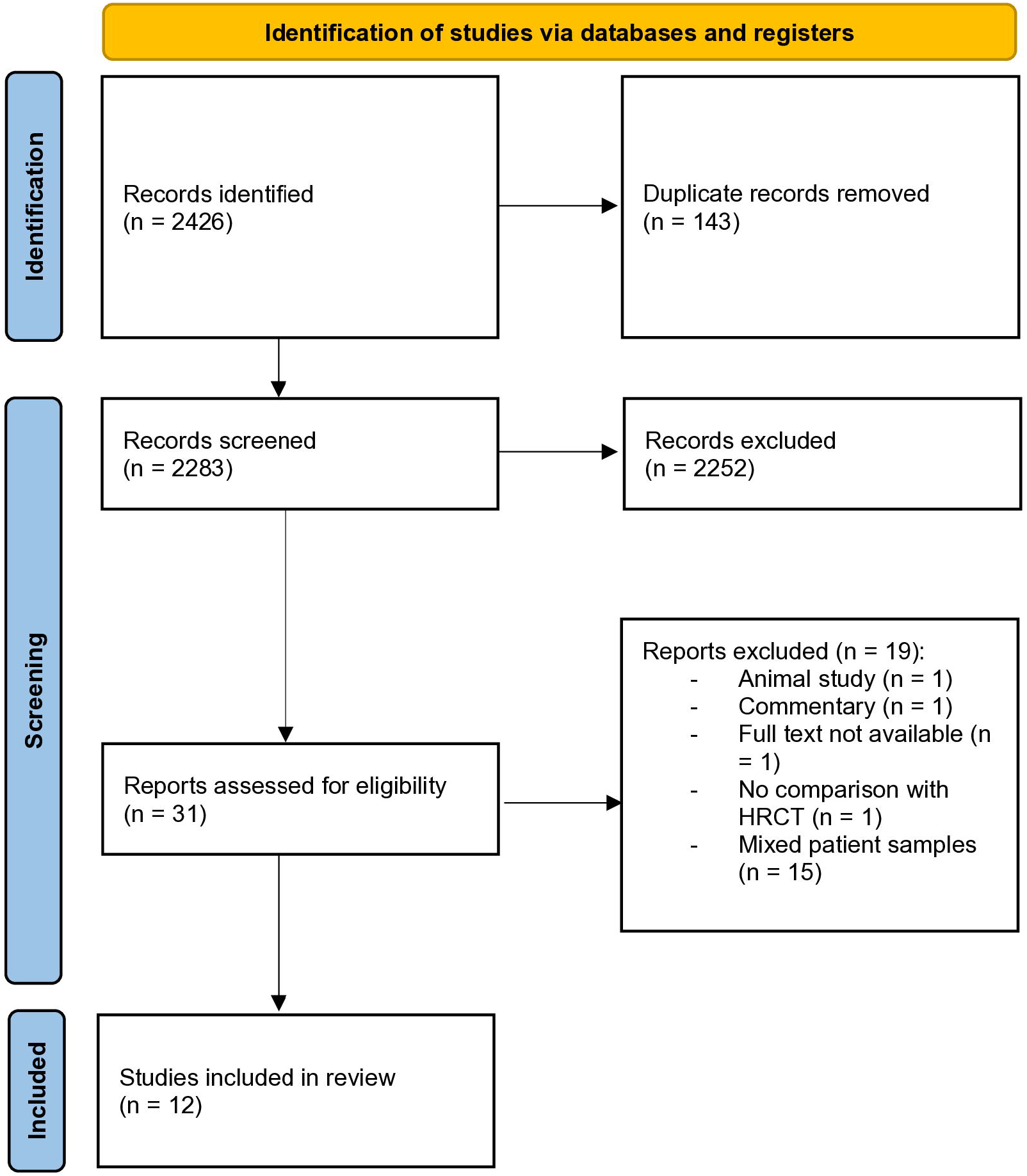

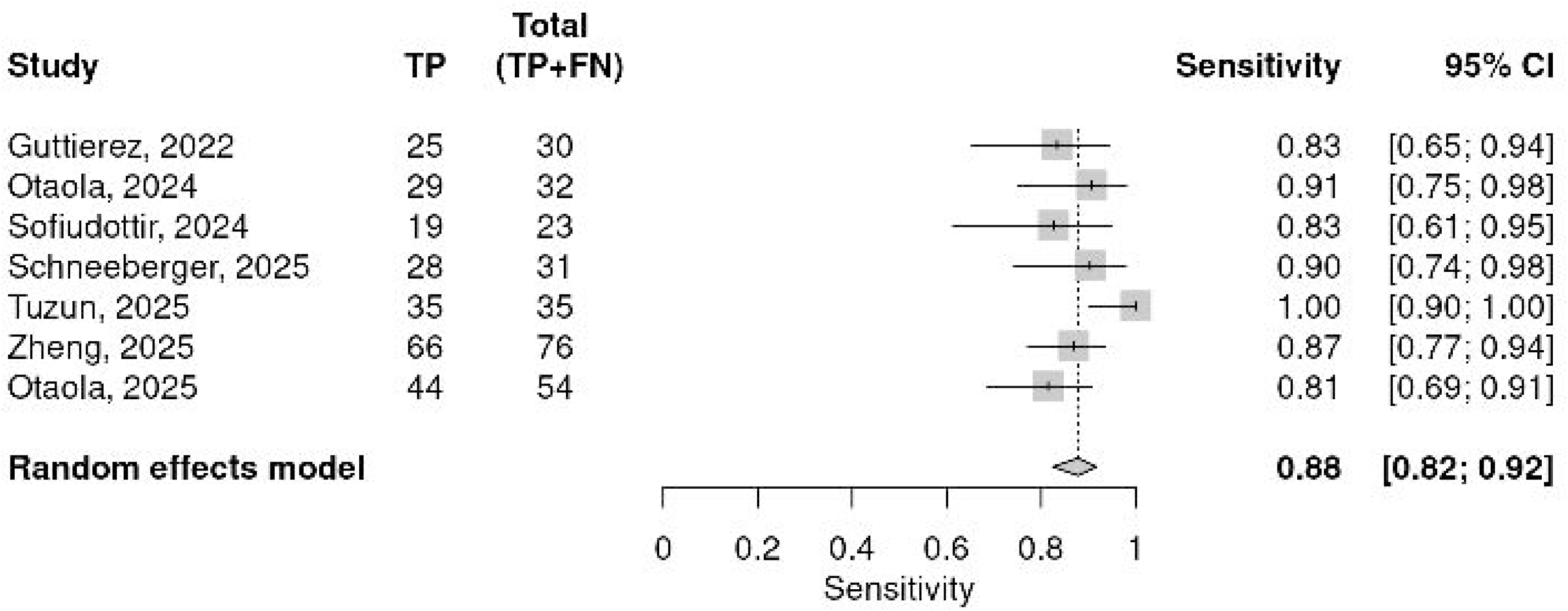

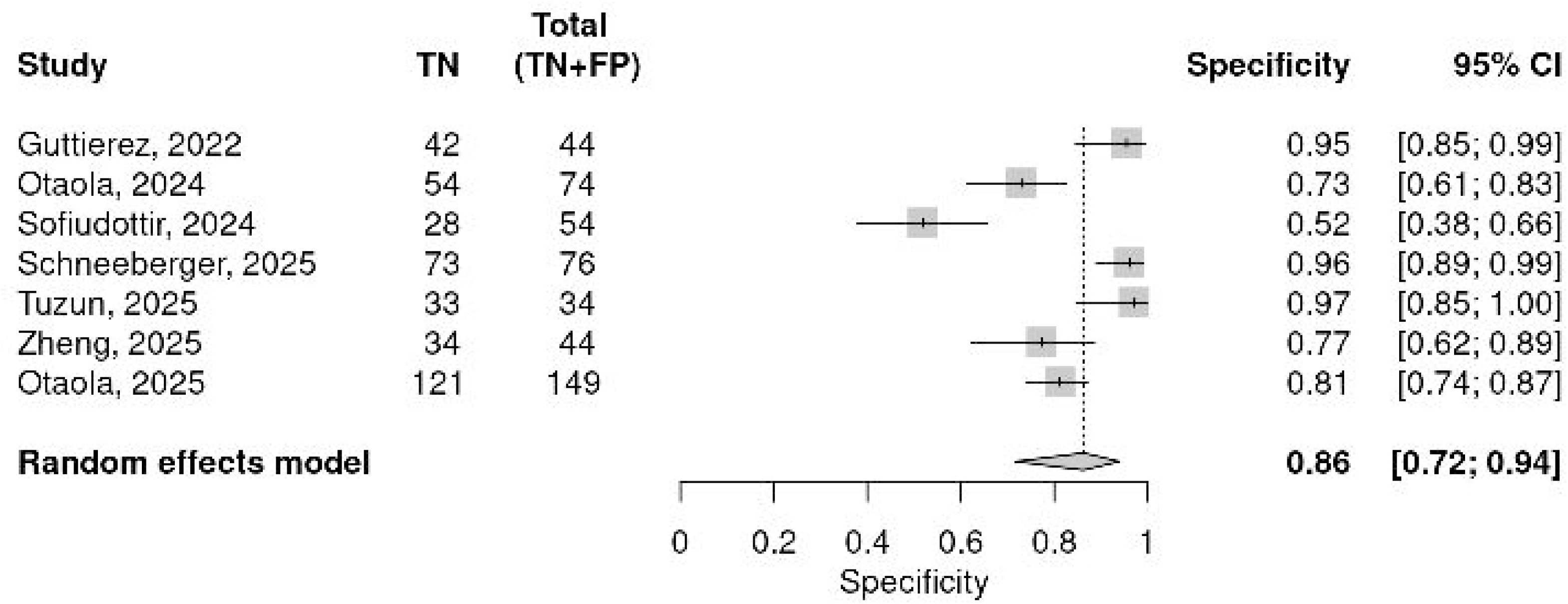

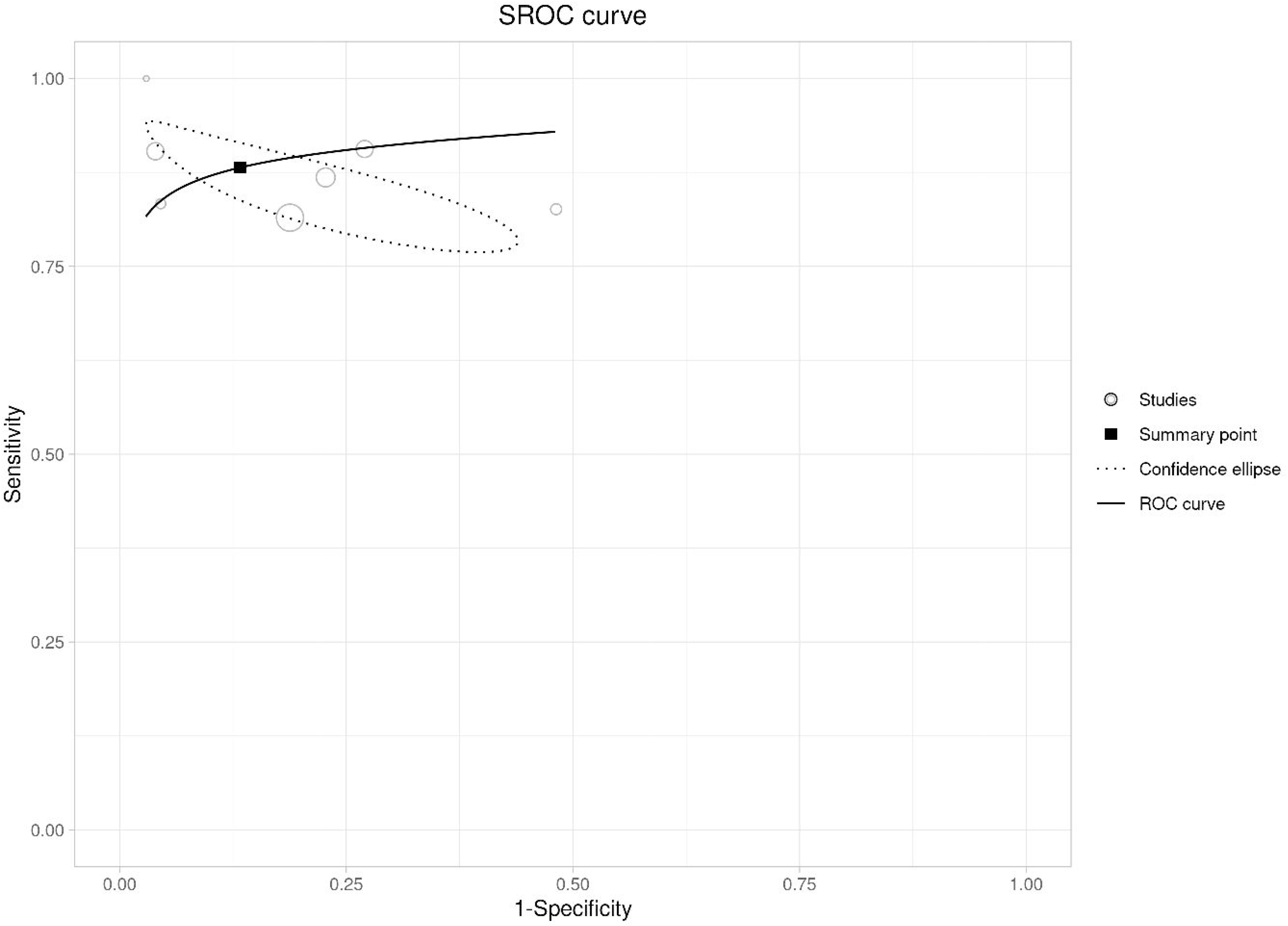

