## Supplemental file 1 for "Screening for rheumatoid arthritis-associated interstitial lung disease using lung ultrasound: a systematic review and meta-analysis of recent studies"

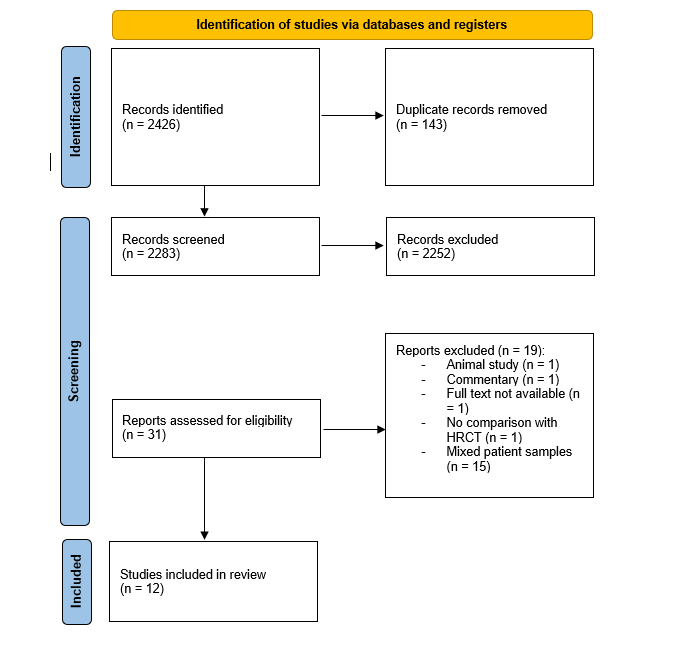

Fig. 1. PRISMA systematic review selection flowchart.

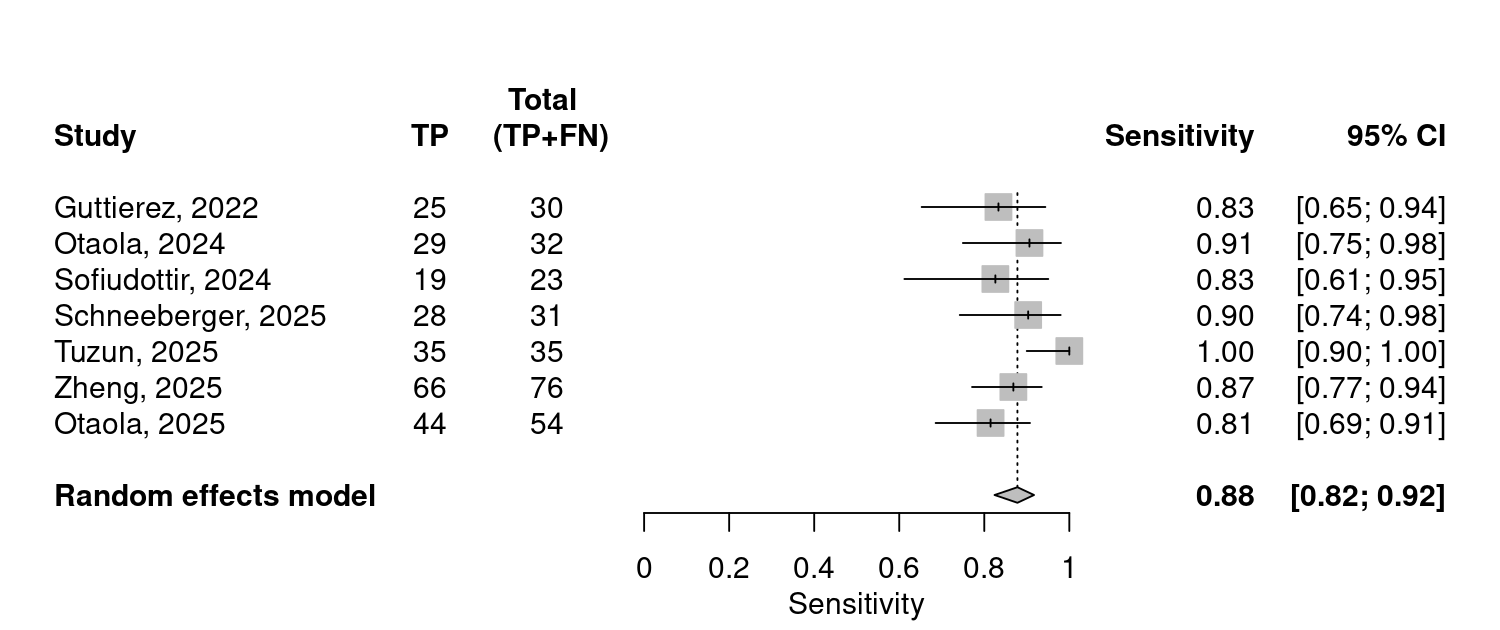

Fig. 2. Forest plot of individual and pooled sensitivity for lung ultrasound in RA-ILD. Random effects model was used due to high heterogeneity.

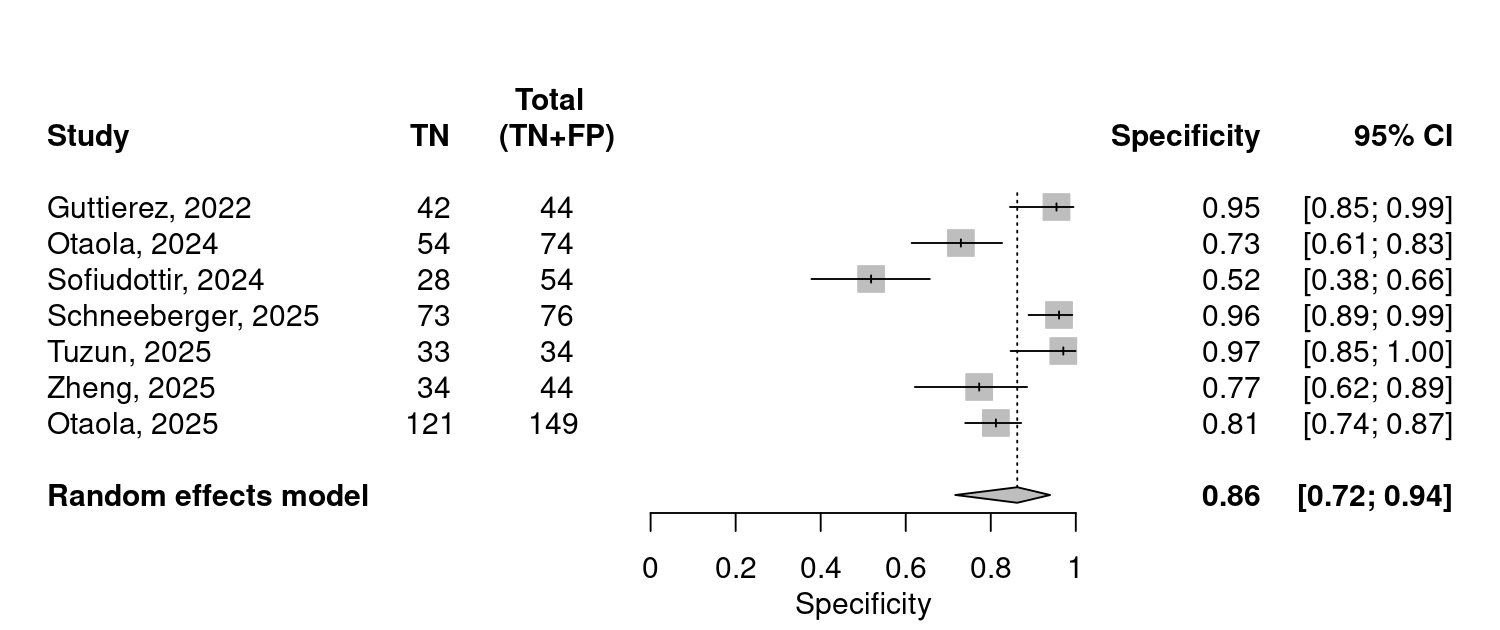

Fig. 3. Forest plot of individual and pooled specificity for lung ultrasound in RA-ILD. Random effects model was used due to high heterogeneity.

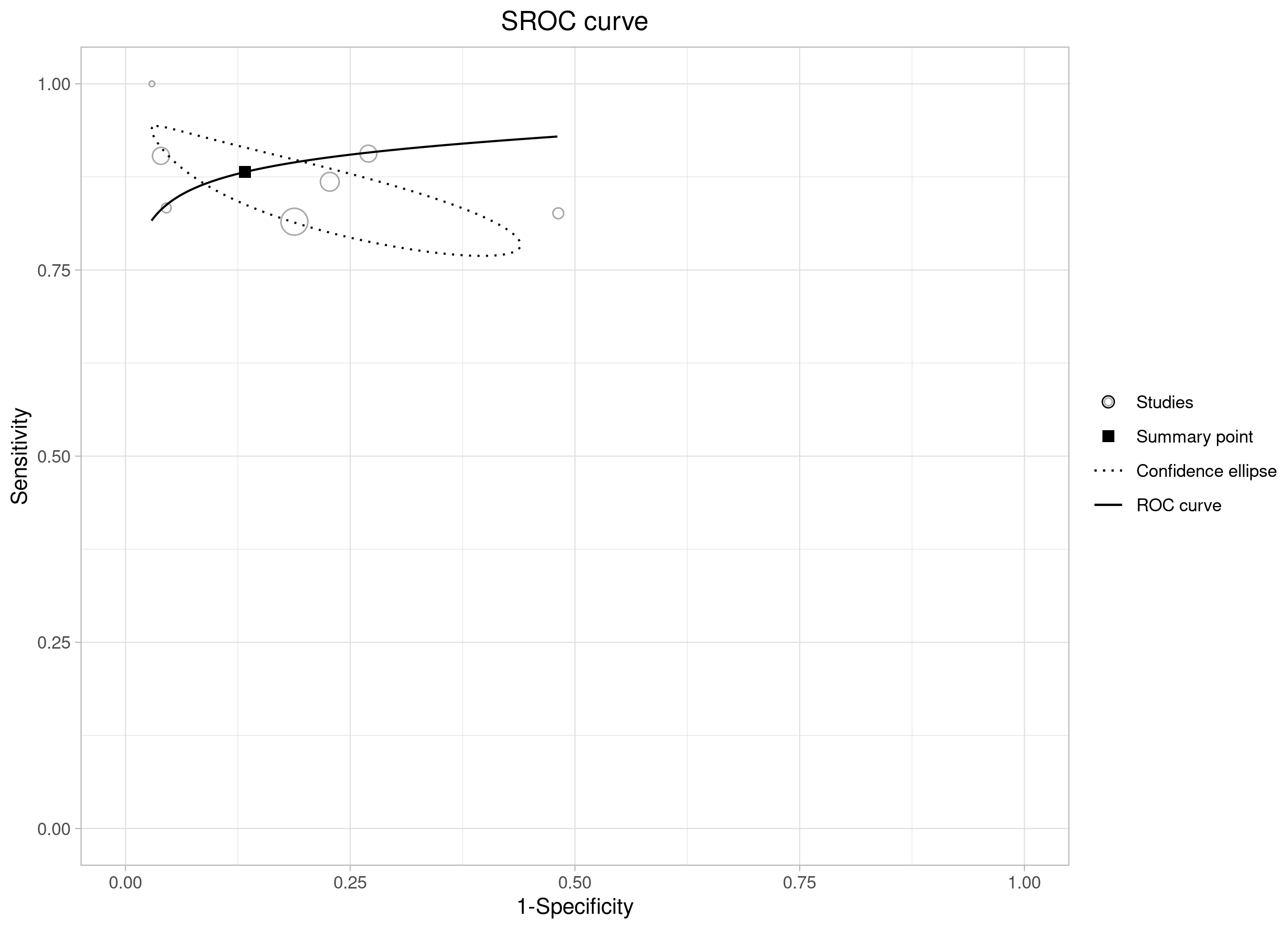

Fig. 4. SROC curve of pooled meta-analysis data.

|  | Estimate |
| --- | --- |
| Var logit(sen) | 0.135 |
| Var logit(spe) | 1.353 |
| MOR sensitivity | 1.419 |
| MOR specificity | 3.033 |
| Bivariate I2 | 0.001 |
| Area 95% Prediction Ellipse | 0.149 |

Heterogeneity of studies included in meta-analysis.
